# Probabilistic seasonal dengue forecasting in Vietnam using superensembles

**DOI:** 10.1101/2020.05.20.20108019

**Authors:** Felipe J Colón-González, Leonardo Soares Bastos, Barbara Hofmann, Alison Hopkin, Quillon Harpham, Tom Crocker, Rosanna Amato, Iacopo Ferrario, Francesca Moschini, Samuel James, Sajni Malde, Eleanor Ainscoe, Vu Sinh Nam, Dang Quang Tan, Nguyen Duc Khoa, Mark Harrison, Gina Tsarouchi, Darren Lumbroso, Oliver Brady, Rachel Lowe

**Affiliations:** Centre for Mathematical Modelling of Infectious Diseases, London School of Hygiene & Tropical Medicine, United Kingdom; HR Wallingford, United Kingdom; Met Office, United Kingdom; National Institute of Hygiene and Epidemiology, Vietnam; General Department of Preventive Medicine, Vietnam; Centre on Climate Change and Planetary Health, London School of Hygiene & Tropical Medicine, London, UK; Barcelona Institute for Global Health (ISGlobal), Barcelona, Spain; Scientific Computing Program, Oswaldo Cruz Foundation (Fiocruz), Rio de Janeiro, Brazil; School of Environmental Sciences, University of East Anglia, Norwich, UK; Tyndall Centre for Climate Change Research, University of East Anglia, Norwich, UK

## Abstract

Timely information is key for decision-making. The ability to predict dengue transmission ahead of time would significantly benefit planners and decision-makers. Dengue is climate-sensitive. Monitoring climate variability could provide advance warning about dengue risk. Multiple dengue early warning systems have been proposed. Often, these systems are based on deterministic models that have limitations for quantifying the probability that a public health event may occur. We introduce an operational seasonal dengue forecasting system where Earth observations and seasonal climate forecasts are used to drive a superensemble of probabilistic dengue models to predict dengue risk up to six months ahead. We demonstrate that the system has skill and relative economic value at multiple forecast horizons, seasons, and locations. The superensemble generated, on average, more accurate forecasts than those obtained from the models used to create it. We argue our system provides a useful tool for the development and deployment of targeted vector control interventions, and a more efficient allocation of resources in Vietnam.

## Introduction

Dengue is a mosquito-transmitted viral infection spread by *Aedes* mosquitoes in urban and periurban environments in tropical and subtropical countries **(*Powell and Tabachnick, 2013; Li et al., 2014; Kraemer et al., 2015*)**. About half of the global population is at risk of dengue transmission **(*Brady et al., 2012; Bhatt et al., 2013*)**. There is no specific antiviral treatment for dengue, and vaccination is restricted to seropositive individuals **(*World Health Organization, 2018*)**. Dengue prevention relies on mosquito control measures which are primarily insecticide-based **(Dusfour *et al., 2019*)**. The increasing resistance to insecticides highlights the need for targeted and effective interventions **(*Moyes et al., 2017*)**.

Vietnam is particularly affected by dengue with an estimated burden of about two million yearly infections **(*Bhatt et al., 2013; Shepard et al., 2016*)**, although, on average, only 95,000 cases have been reported annually to the Ministry of Health over the period 2002-2020. The economic impact of dengue in Vietnam is estimated to be USD 30-95 million per annum **(*Shepard et al., 2014,2016; Hunget al., 2018*)**. Dengue in Vietnam is primarily spread by *Ae. aegypti* and, to a lesser extent, by ***Ae. albpoictus* (*Tsunoda et al., 2014; Thi et al., 2017*)**.

In Vietnam, dengue is characterized by strong seasonality and substantial inter-annual and spatial variability (Supplementary Figure 1). Dengue exhibits different behaviour in different parts of the country. In the north, where temperatures are lower than in the rest of the country, most provinces have few or no cases. An exception is Hanoi, which has reported, on average, about 8,700 dengue cases per year over the past ten years. In central and southern provinces where temperatures are warmer, many provinces report thousands of cases annually albeit with large interannual variation.

Dengue control in the country primarily involves community engagement and mobilisation to reduce breeding sites, and outdoor low-volume insecticide spraying in the vicinity of reported dengue cases to kill adult mosquitoes **(*Cuong et al., 2013*)**. One limitation of dengue control measures is that they are essentially reactive, meaning they take place after cases have occurred. This situation hampers the ability of public health professionals to reduce the magnitude and severity of outbreaks. Dengue surveillance is mostly passive, relying on clinical cases reported by patients seeking healthcare **(*Cuong et al., 2013*)**. The diagnosis and reporting of dengue cases typically suffers delays which vary across time and space **(*Bastos et al., 2019*)**. This situation hinders the timely generation and communication of information on when and where transmission occurs, limiting the ability of health professionals to plan and execute control measures.

If accurate predictions are available, public health decision-makers and planners could design, implement and target interventions to the most at-risk places in a timely fashion. Disease models driven by Earth observations have been valuable for predicting dengue risk ahead of time, supporting decision-making in multiple settings **(*Lowe et al., 2016, 2017; Colón-González et al., 2018a*)**. Climate variation is one of the main drivers of dengue ecology. Temperature, for example, regulates the development, biting, and reproduction rates, and the spatial distribution of *Aedes* mosquitoes **(*Gage et al., 2008; Reinhold et al., 2018; Kraemer et al., 2019*)**. The concentration and replication of dengue viruses within mosquitoes is also temperature dependent (***Watts et al., 1987; Gage et al., 2008*)**. Rising temperatures increase dengue transmission to an optimum range of 26–29°C **(*Mordecai et al., 2017*)**. Large diurnal temperature ranges (> 20°C) reduce transmission and increase mosquito mortality **(*Lambrechts et al., 2011*)**. Precipitation modulates the creation or flush away of mosquito breeding sites **(*Stewart Ibarra et al., 2013*)**. Rising humidity increases dengue risk **(*Colón-González et al., 2017*)** as relative humidity levels of at least 50-55% prolong mosquito survival **(*Simon-Oke and Olofintoye, 2015*)**. Wind speed reduces the biting activity of mosquitoes reducing dengue risk **(*Sedda et al., 2018*)**. Delayed effects of climate on dengue have been demonstrated and correspond to climatic influences on dengue through their effect on the life cycle of both mosquitoes and the dengue virus **(*Naish et al., 2014*)**. Consequently, the climatic conditions during the low-dengue season may be indicative of dengue incidence in the following high-dengue season **(*Lauer et al., 2018; Lowe et al., 2018*)**.

Multiple studies have highlighted the potential usefulness of seasonal-climate-driven epidemiological surveillance for decision-making and planning **(*Lowe et al., 2014; Lauer et al., 2018; Tompkins and Di Giuseppe, 2015; Tompkins et al., 2019*)**. These studies have used subseasonal (i.e. between two weeks and two months ahead) forecasts to inform disease models, and compute predictions of dengue risk. There has been limited progress in using subseasonal-to-seasonal climate forecasting to compute prospective forecasts on a routine basis. There are several challenges for implementing operational and sustainable subseasonal (henceforth seasonal) early warning systems **(*Thomson et al., 2014*)**. Some of these challenges include the lack of multi-decadal health data sets with which to train and validate seasonal-climate-driven early warning systems, the common mismatch of scales between climate data outputs and data used for decision-making, and a general lack of consensus as to how to communicate uncertainties to users **(*Tompkins et al., 2019*)**.

Dengue early warning systems driven by Earth observations and seasonal climate forecasts have been proposed using a range of modelling approaches **(*Yamana et al., 2016*)**, including autoregressive integrative moving average (ARIMA) **(*Eastin et al., 2014*)**, deterministic regression **(*Hii et al., 2013; Lauer et al., 2018*)**, spatio-temporal Bayesian hierarchical models **(*Lowe et al., 2016, 2017*)**, least absolute shrinkage and selection operator (LASSO) regression **(*Chen et al., 2018*)**, and machine learning **(*Stolerman et al., 2019*)**. Often, models are validated using block cross-validation to select the model specification with the lowest out-of-sample predictive error **(*Lowe et al., 2016; Lauer et al., 2018*)**. This approach takes advantage of all available data to make repeated out-of-sample model predictions, which increases the robustness of model adequacy and skill score statistics. One drawback of this method is that it does not preserve the time ordering of the data. Also, predictions are computed for some time periods using a model trained on data from a later time period **(*Bergmeir and Benítez, 2012*)**.

Previous dengue risk prediction studies have relied on outputs from one or two competing models (e.g. ***Lowe et al., 2016; Lauer et al., 2018*)**. However, combining forecasts from multiple competing models into a superensemble can result in more accurate predictions than those from any individual model (see, for example, ***Yamana et al., 2016; Johansson et al., 2019*)**. The use of model superensembles for the development of dengue early warning systems has been seldom explored. Moreover, predictions are typically made for a selected year or month **(*Lowe et al., 2014, 2016; Lauer et al., 2018*)** rather than for a series of lead times into the future, or a whole season **(*Lowe et al., 2017*)**. In some cases, systems are designed exclusively for research purposes in isolation from relevant stakeholders who may become potential users.

Typically, dengue early warning systems are based on deterministic models **(*Hii et al., 2013; Eastin et al., 2014; Lauer et al., 2018*)** which may under-represent heterogeneity and stochastic cessation of transmission. However, decision-makers are increasingly interested in understanding the uncertainties related to the models used to develop decision-support tools, and in the probabilities that an event of public health concern may or may not take place **(*Lowe et al., 2014; Colón-González et al., 2018b; Lake et al., 2019*)**. Spatio-temporal probabilistic models have the advantage of being able to quantify the probability that an event (e.g. an outbreak) may occur at specific times and for specific locations. Public health officials may be more inclined to take action if the probability of observing an outbreak exceeds a certain value **(*Lowe et al., 2016*)**. Both modellers and decision-makers should pay attention to, and agree on the definition of outbreaks thresholds so that model predictions are a useful guide for planning and decision-making.

In several countries, including Vietnam, outbreaks are defined using a so called ***endemic channel* (*Badurdeen et al., 2013; Runge Ranzinger et al., 2014*)** which corresponds to the mean number of cases per month or season over a long-term period **(*Brady et al., 2015*)**. In Vietnam, endemic channels are defined for each province using the last five years of dengue surveillance data. Outbreak years are removed from the computation of the endemic threshold. When dengue cases exceed the mean plus two standard deviations, an *outbreak* is declared. One limitation of this approach is that often outbreak years are removed arbitrarily or quasi-quantitatively to increase the sensitivity of the outbreak threshold **(*Brady et al., 2015*)**. In areas where dengue incidence is typically low (e.g. < 10 cases per month), the endemic channel may be frequently exceeded generating statistical alarms of little public health importance **(*Noufaily et al., 2019*)**. Despite these limitations, endemic channels are widely used for dengue control decision-making in a variety of countries and provide a practical decision point around which forecasts can be targeted **(*Hussain-Alkhateeb et al., 2018; Olliaro et al., 2018*)**.

Here, we introduce a superensemble of probabilistic spatio-temporal hierarchical dengue models driven by Earth observations and seasonal climate forecasts. The model framework was codesigned with stakeholders from the World Health Organization, the United Nations Development Programme, the Vietnamese Ministry of Health, the Pasteur Institute Ho Chi Minh City, the Pasteur Institute Nha Trang, the Institute of Hygiene and Epidemiology Tay Nguyen (TIHE), and the National Institute of Hygiene and Epidemiology (NI HE). The system is designed to generate monthly estimates of dengue risk across Vietnam (331,210 km^2^) at the province level (*n* = 63) in *near-real time* (i.e. current time minus processing time). The superensemble is used in an expanding window time series cross-validation framework **(*Hyndman and Athanasopoulos, 2014*)** to generate probabilistic dengue forecasts, which allow us to calculate the probability of exceeding pre-defined dengue outbreak thresholds for a forecast horizon (i.e. lead time) of one to six months.

The superensemble constitutes the dengue fever component of a forecasting system called *D-MOSS* (i.e. Dengue forecasting MOdel Satellite-based System. The system operates using a suite of Earth observation data sources from satellites, and has its first implementation in Vietnam. Vietnam is divided into 63 provinces which are subdivided into 713 districts and has an estimated population of 95.5 million people. The D-MOSS system produces results at the province level and is accompanied by a skill assessment that is applied consistently across the whole of Vietnam. The intention is to give an overall evaluation of the performance of our method, rather than an assessment that pertains to the characteristics of any particular province.

## Results

We fitted a total of 14,592 different models (128 unique model specifications across 114 forecast months, i.e. January 2007 to December 2016). Dengue data was modelled using Earth observations of mean monthly values of minimum temperature, maximum temperature, precipitation amount per day, specific humidity, diurnal temperature range, wind speed, and sea surface temperature anomalies for the Niño 3.4 Region (i.e. 5°S–5°N and 170°–120°W) as explanatory variables. Each variable was included in isolation, as well as with all possible combinations (see Methods and Materials). Models included the proportion of people living in urban, periurban and rural areas (henceforth land-cover data). Land-cover data were included in all models as they change annually and not monthly (see Methods and Materials). Predictions up to six months ahead were made at each time step starting in January 2007 using seasonal climate hindcasts (retrospective forecasts) data from the UK Met Office Global Seasonal Forecasting System version 5 **(*MacLachlan et al., 2015; Scaife et al., 2014*)**.

### Model superensemble

Models were evaluated against five verification metrics: continuous rank probability score (CRPS), root mean squared error (RMSE), mean absolute error (MAE), deviance information criterion (DIC), and Watanabe-Akaike information criterion (WAIC). We selected the best two models within each verification metric to create a model superensemble. Verification metrics were computed for each model specification at each time step and averaged across the whole time series. For all verification metrics, lower values indicate a smaller difference between the forecasts and the observations. The five models with the lowest values for the selected verification metrics (i.e. best performing) were selected to generate a superensemble (Table 1). Combining all best performing models into a model superensemble led to a lower CRPS and MAE than any of the five competing models. The RMSE of the superensemble, however, was higher than two of the five models.

When stratified by forecast horizon time, the predictive ability of the superensemble was greater than or comparable to that of the best performing competing models (Figure 1). Across all metrics, the predictive ability of the superensemble deteriorated as the forecast horizon increased from one to six months ahead. This situation is also evident where the forecast ensemble mean and its 95% credible interval (i.e. the interval in the domain of the posterior probability distribution) are plotted against the observed number of dengue cases (Supplementary Figure 2). Notice that the accuracy of the predictions worsens as the forecast horizon expands. We noted that the credible intervals of the predictions gradually became narrower as the number of months used to train the models increased (Supplementary Figure 2).

**Table 1.**
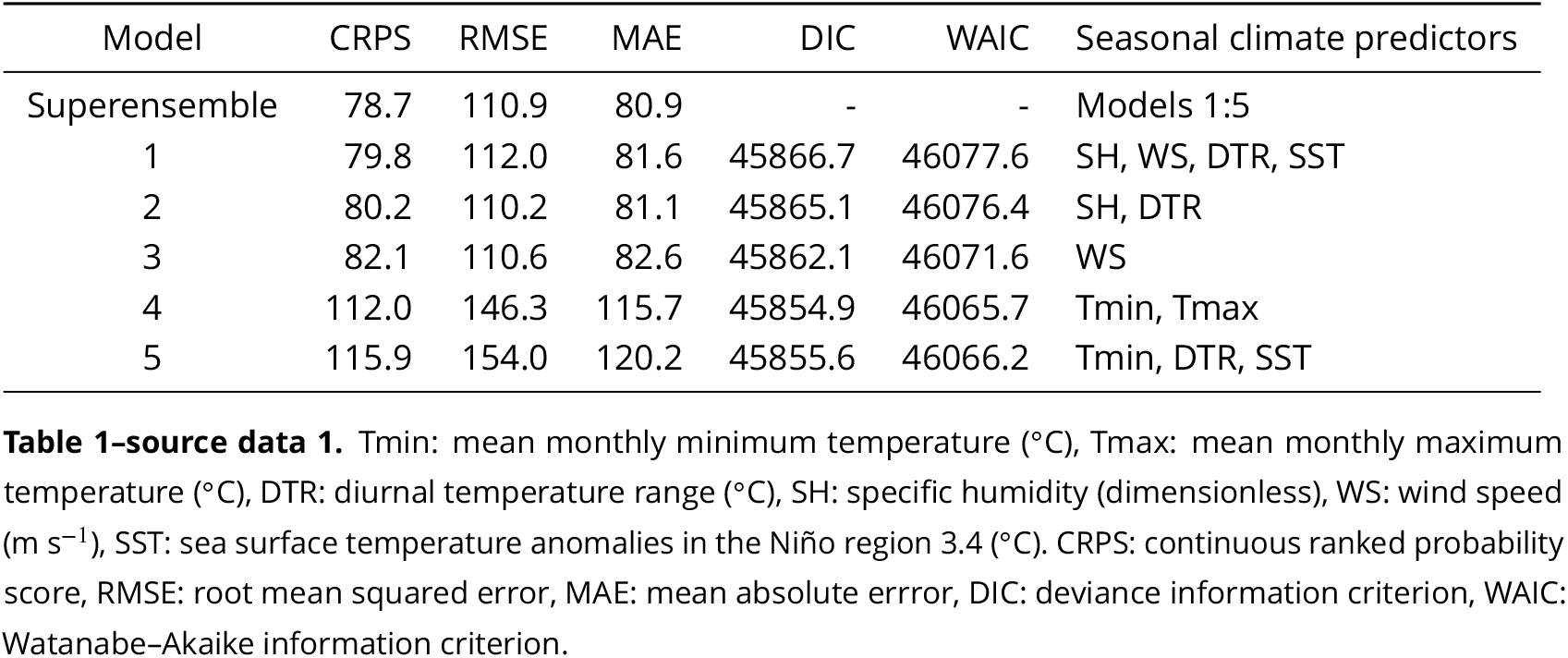
Verification metrics and seasonal climate predictors of the model superensemble, and the best performing models.

source data 1. Tmin: mean monthly minimum temperature (°C), Tmax: mean monthly maximum temperature (°C), DTR: diurnal temperature range (°C), SH: specific humidity (dimensionless), WS: wind speed ( $\text{m s}^{-1}$ ), SST: sea surface temperature anomalies in the Niño region 3.4 (°C).
| Model | CRPS | RMSE | MAE | DIC | WAIC | Seasonal climate predictors |
| --- | --- | --- | --- | --- | --- | --- |
| Superensemble | 78.7 | 110.9 | 80.9 | - | - | Models 1:5 |
| 1 | 79.8 | 112.0 | 81.6 | 45866.7 | 46077.6 | SH, WS, DTR, SST |
| 2 | 80.2 | 110.2 | 81.1 | 45865.1 | 46076.4 | SH, DTR |
| 3 | 82.1 | 110.6 | 82.6 | 45862.1 | 46071.6 | WS |
| 4 | 112.0 | 146.3 | 115.7 | 45854.9 | 46065.7 | Tmin, Tmax |
| 5 | 115.9 | 154.0 | 120.2 | 45855.6 | 46066.2 | Tmin, DTR, SST |

**Figure 1.**
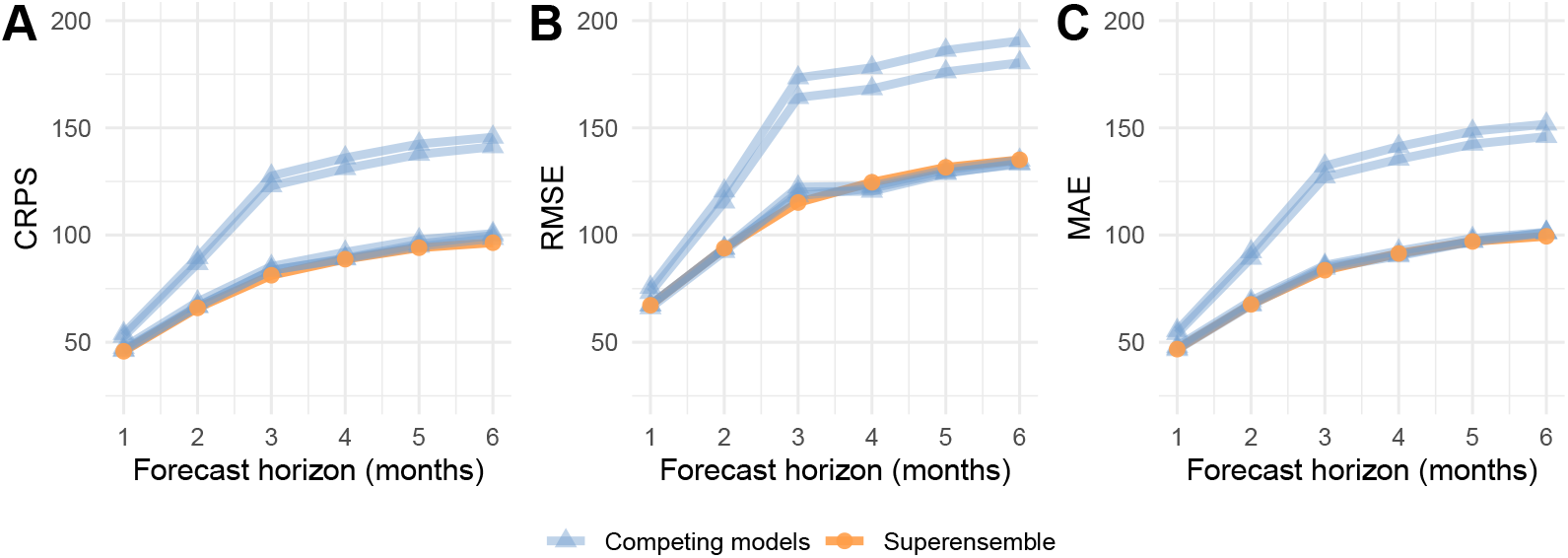
Forecasting metrics of five competing models (blue) compared against a model superensemble (orange). Metrics shown are (A) continuous rank probability score (CRPS), (B) root mean squared error (RMSE), and (C) mean absolute error (MAE). All metrics are shown as a function of the forecast horizon.

The predictive ability of the model also varied with the month of the year (Figure 2). Overall, larger discrepancies between observed and predicted values were observed between July and December, when typically more cases are reported. Similar patterns were observed for the CRPS, RMSE and MAE (Supplementary Figure 3).

The skill of the forecast showed significant spatiotemporal variation. Skill was evaluated using the continuous rank probability skill score, CRPSS **(*Bradley and Schwartz, 2011*)** comparing the CRPS of the superensemble to that of a baseline model (see Methods and Materials). CRPSS values larger than zero indicate a better predictive skill than that of the baseline model. Figure 2 shows that the CRPSS was consistently better than the baseline across the whole country for the period December to July. From August to November the skill is reduced for selected provinces in the central and southern regions characterised by larger dengue incidence variability.

Supplementary Figure 4 shows the observed and posterior predictive mean dengue cases across the 63 provinces computed one month ahead using the model superensemble. The model superensemble is able to reproduce the spatiotemporal dynamics of dengue fever with reasonable skill although predictions tend to underestimate the number of cases particularly during large outbreaks.

**Figure 2.**
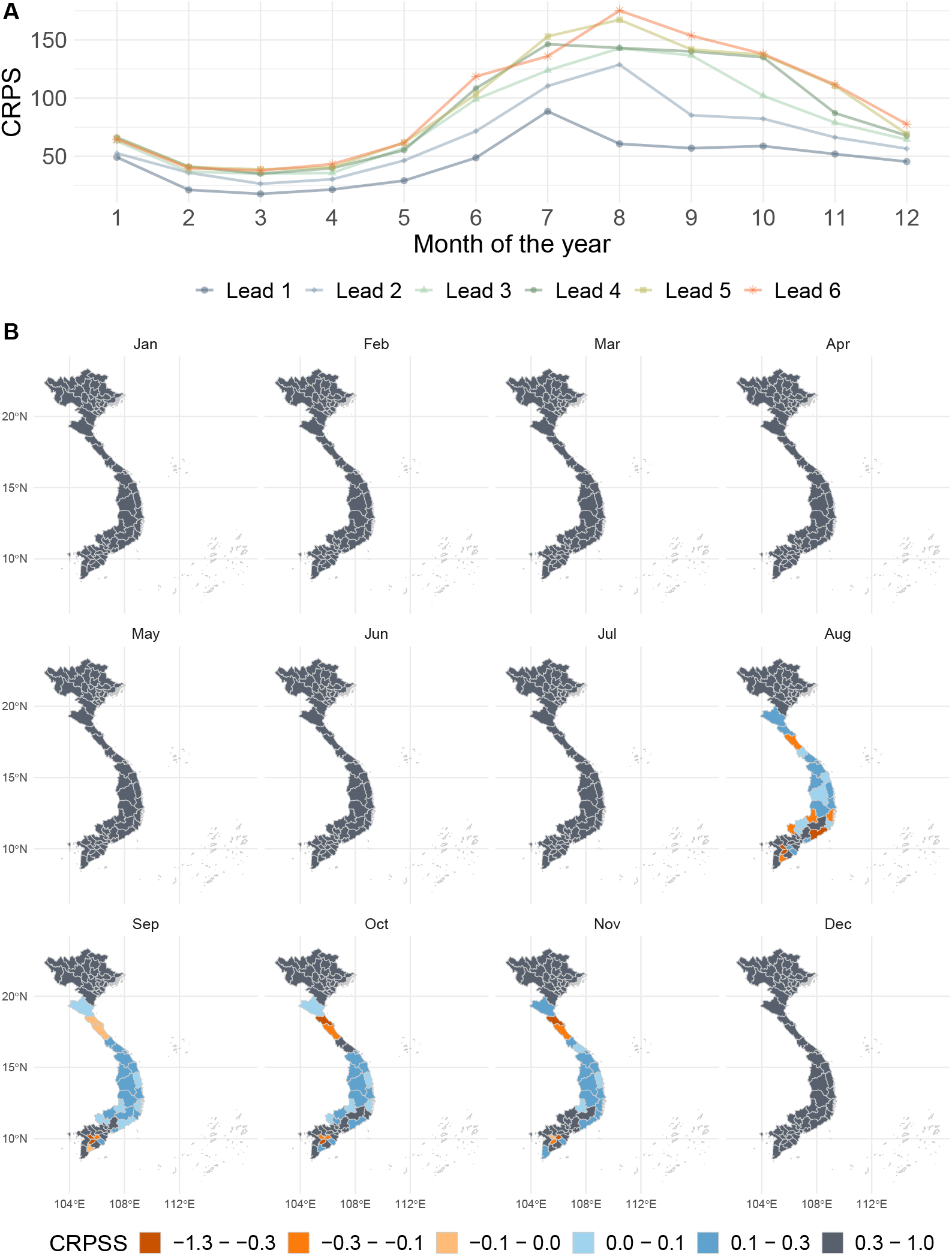
(A) Continuous rank probability score (CRPS) of the model superensemble arranged by month and forecast horizon, averaged across Vietnam. (B) Spatiotemporal variation of the continuous rank probability skill score (CRPSS) of the model superensemble. Gray and blue shaded areas indicate a better performance of the superensemble compared to a baseline model. Orange areas indicate a lower performance of the superensemble compared to the baseline model.

### Outbreak detection

The skill of the model for outbreak detection was evaluated using the Brier score **(*Brier, 1950*)**. For the Brier score, smaller values are better. Four moving outbreak thresholds were defined to evaluate the skill of the superensemble for outbreak prediction: i) the endemic channel plus one standard deviation, ii) the endemic channel plus two standard deviations, iii) the *75^th^* percentile of the distribution of dengue cases, and iv) the 95*^th^* percentile of the distribution of dengue cases. The endemic channel was calculated as the number of dengue cases per month and per province over the previous five years in agreement with current practice at the Vietnamese Ministry of Health. The 75*^th^* and 95*^th^* percentiles were calculated over the whole observational period at each time step (see Methods and Materials). We then calculated the probability of exceeding the moving outbreak threshold based on the posterior marginal distribution of the predicted number of cases.

Predictive skill was evaluated by comparing the predicted probability of exceeding the moving outbreak threshold to observed outbreaks. Observed outbreak months were defined as months where the number of recorded dengue cases exceeded the moving outbreak thresholds. Figure 3 shows the observed outbreaks and their corresponding predicted probabilities based on the marginal posterior distribution of the model output. Figure 4 depicts the outbreak detection skill of the superensemble across the forecast horizon (top) and time of the year (bottom).

As expected, the highest skill was achieved at a lead time of one month, after which the skill of the superensemble gradually declines. Across the forecast horizon, the highest skill was observed when using an outbreak threshold based on the 95*^th^* percentile of the distribution of dengue cases, followed by the endemic channel plus two standard deviations. Stratified by month of the year, the skill of the superensemble was generally greater between April and October. We noted that using the endemic plus one or two standard deviations results in a significant decrease in skill between June and July. This situation is not observed when using percentiles. When stratified by month, skill was larger using the 95*^th^* percentile of the distribution of dengue cases followed by the endemic channel plus two standard deviations.

There was significant spatial variation in the outbreak detection skill of the superensemble. Figure 5 shows that across all moving outbreak thresholds, skill was greater in the northern provinces compared to the central and southern provinces. Note that using the 95*^th^* percentile as a threshold results in a larger number of provinces with a low Brier score (< 0.1).

Public health officials may be more likely to take preventive action if the probability of observing an outbreak exceeds a certain value **(*Lowe et al., 2016*)**. We investigated different probability thresholds to define the optimal cut-off value that maximised the sensitivity and specificity of the superensemble. As a metric of the accuracy of the forecasts, we computed the area under the receiver operating characteristic curve (AUC) where an AUC value of 1 represents perfect skill, and a value of 0.5 represents no better skill than a random guess. Supplementary Figure 5 indicates that the forecasts always performed better than randomly guessing. As expected, the AUC decreased as the forecast horizon increased. The AUC values ranged between 0.91 and 0.72.

### Relative economic value

A cost-loss analysis of the *relative economic value* of the superensemble was undertaken using the value index **(*Richardson, 2000; Thornes and Stephenson, 2001*)**. The value index ranges between zero and one, with one indicating a perfect forecast. The relative economic value of the superensemble can be interpreted as the cost of using the system relative to the cost of either never preventing outbreaks or always taking action. Location- and time-specific costs and losses may be difficult to quantify. Therefore, our figures are only illustrative.

We defined a range of theoretical epidemic thresholds ranging between the 51*^st^* and the 99*^th^* percentiles of the distribution of dengue cases for the whole time series. Outbreak thresholds were province- and month-specific. Figure 6 shows that the superensemble has a theoretical relative economic value in multiple provinces across a range of cost-loss ratios (i.e. the ratio of the expenses derived of taking preventive action to the potential losses averted) and outbreak thresholds. As the outbreak threshold increases and outbreaks become rarer, the superensemble has relative economic value at lower cost-loss ratios. Overall, larger values were observed when the cost-loss ratio was between 0.2 and 0.3 (i.e. the cost of taking preventive action is between 1/3 and 1/5 of the potential losses caused by not taking action). Supplementary Figure 6 shows the spatiotemporal variation in relative economic value across Vietnam for the forecast horizon of one to six months. Note, the superensemble has relative economic value in areas where dengue is typically endemic such as the central and southern provinces. The superensemble had no relative economic value for northern provinces where dengue is typically absent. The relative economic value gradually declined from an average value of 0.31 one month ahead across 75% (*n*=41) of the provinces, to an average value of 0.13 six months ahead across 62% (*n*=39) of the provinces.

**Figure 3.**
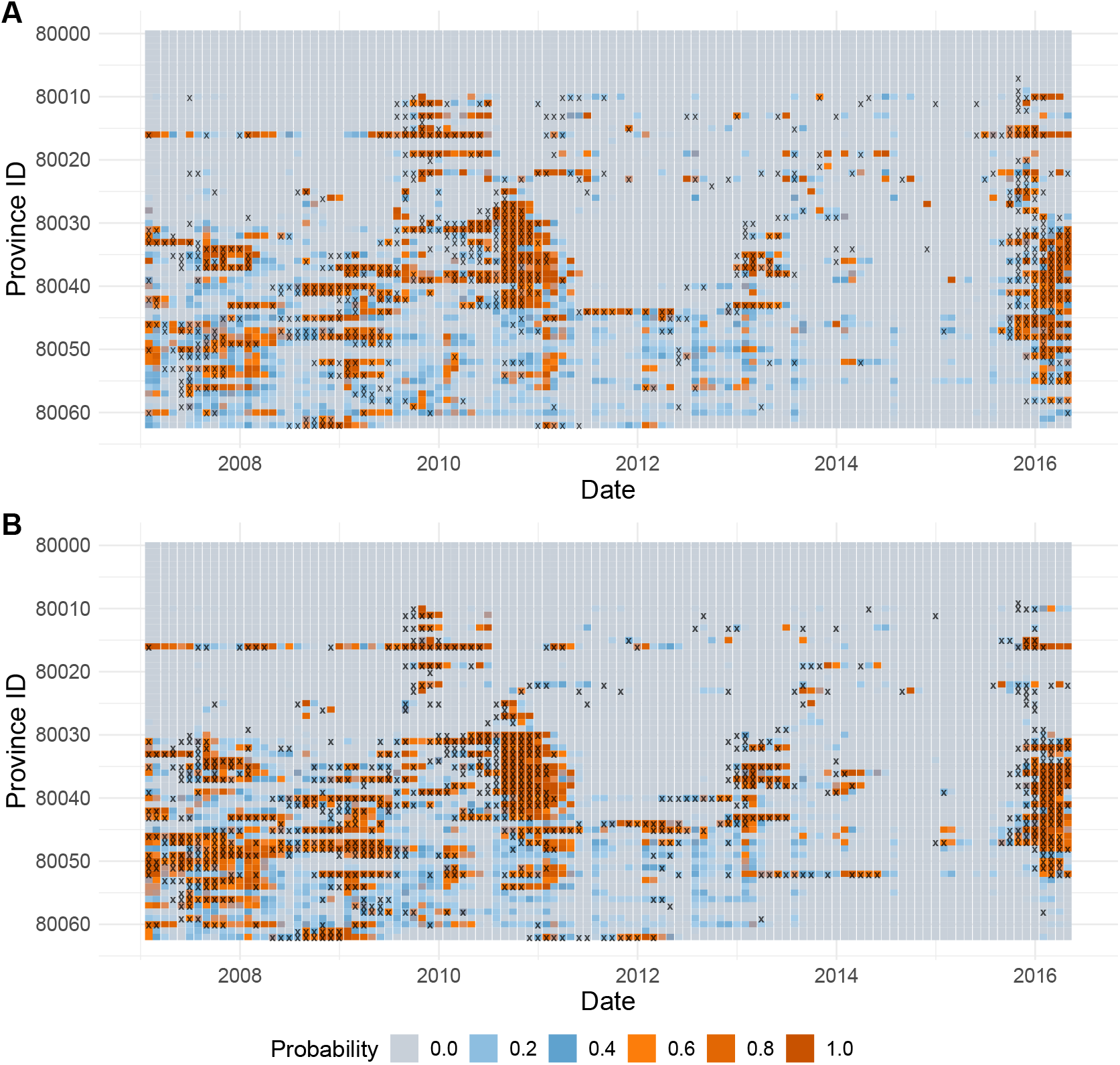
Predicted probability of exceeding the (A) mean plus two standard deviations, and (B) 95*^th^* percentile moving outbreak thresholds one month ahead. The *X* axis represents each of the forecast months. The *Y* axis indicates each of the provinces arranged from north (top) to south (bottom). Observed outbreaks (i.e. observed dengue cases above the outbreak threshold) are marked with a cross. Darker red colours represent a higher probability of exceeding the outbreak threshold.

**Figure 4.**
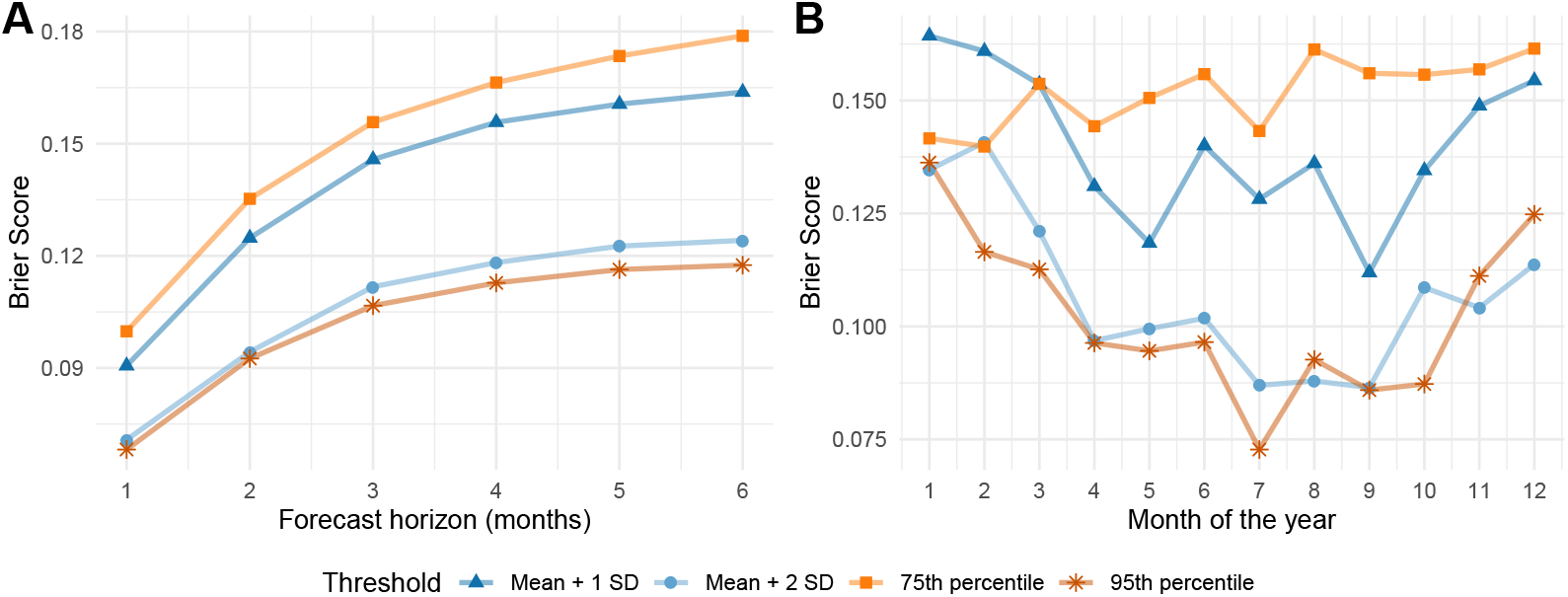
(A) Variation in the Brier score of the superensemble predictions for a forecast horizon (lead time) from one to six months calculated for four different outbreak thresholds. (B) Variation in the Brier score of the superensemble predictions per calendar month, and for four different outbreak thresholds. Scores correspond to their mean values for the period January 2007 to December 2016 (*n* = 114 months), and across 63 Vietnamese provinces. Lower scores indicate a greater accuracy for detecting outbreaks.

**Figure 5.**
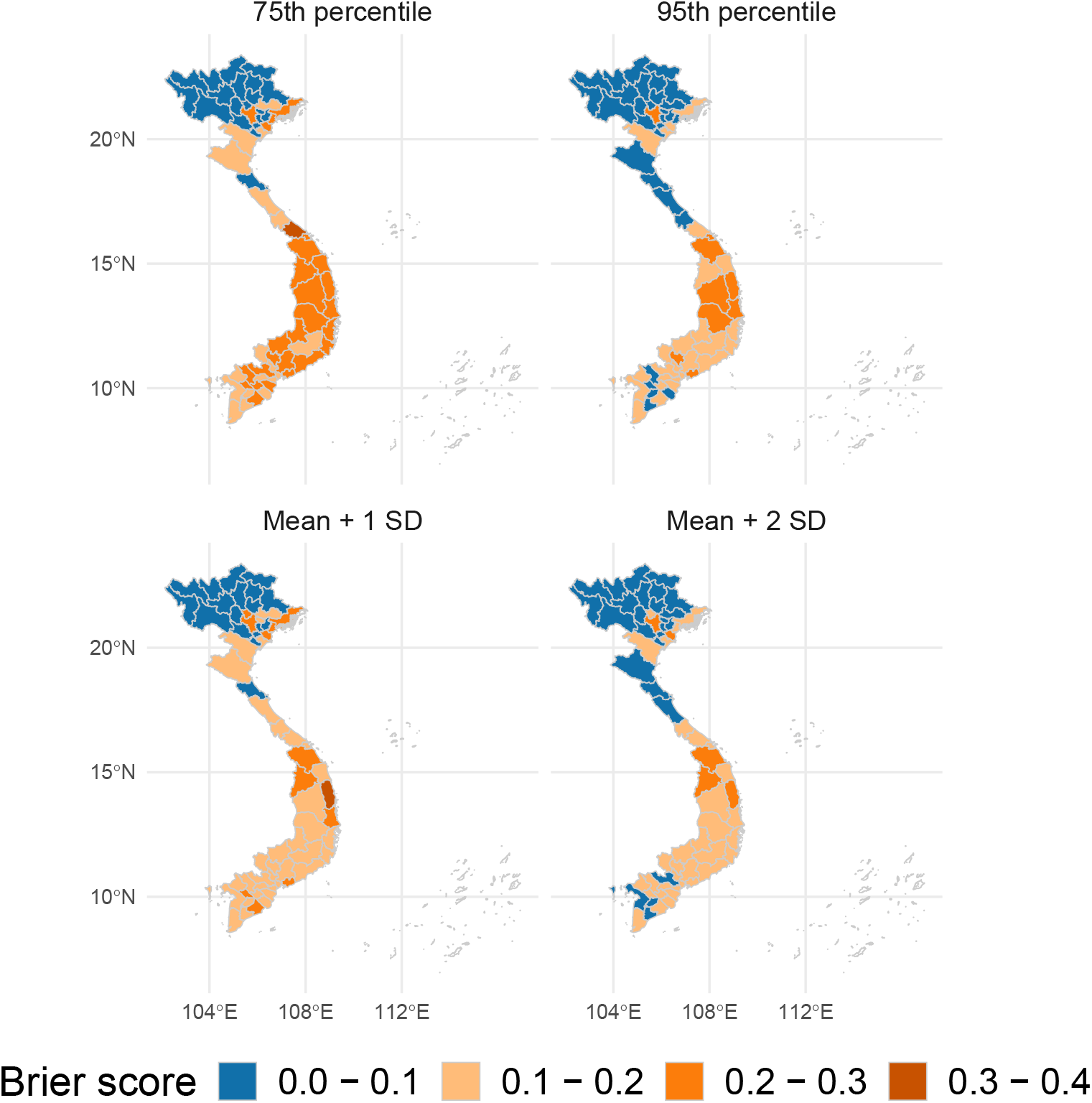
Spatial variation of the mean Brier score including all forecast horizons (lead times) from one to six months calculated for four different outbreak thresholds for the period January 2007 to December 2016 (*n* = 114) for each of the 63 Vietnamese provinces. Lower Brier scores (in blue) indicate a greater accuracy for detecting outbreaks.

**Figure 6.**
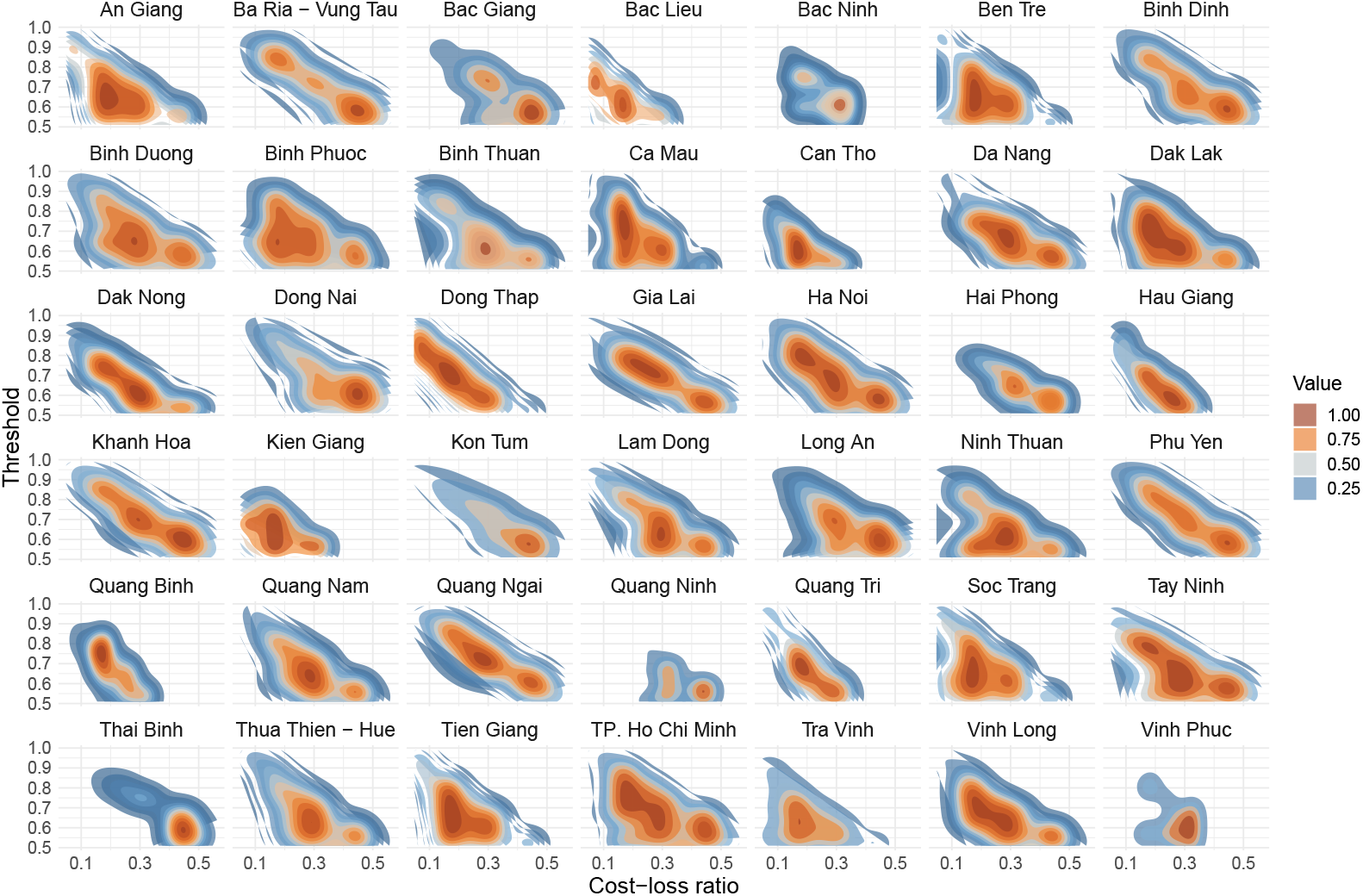
Relative economic value of using the dengue forecasting superensemble one month ahead under a range of cost-loss ratios (*X* axis) and outbreak thresholds (*Y* axis) for 42 Vietnamese provinces where the superensemble showed value at for multiple cost-loss ratios. Orange colours indicate a greater economic value, and blue colours a lower value.

### Prospective predictions of dengue risk

The model superensemble was driven by seasonal climate forecasts to generate dengue forecasts for the period April to September 2020 using near-real time seasonal climate forecast data from the UK Met Office’s Global Seasonal forecasting system version 5 (GloSea 5) **(*MacLachlan et al., 2015; Scaife et al., 2014*)**. The posterior mean of the predicted values for each ensemble member are shown on Figure 7 (solid lines) for all Vietnamese provinces along with the corresponding 95% credible interval (dashed lines). The upper bound of the shaded areas represent percentile-based moving outbreak thresholds computed using observed dengue case data over the period August 2002 to March 2020.

There is little spread between the 42 ensemble members indicating little between-member variability. It is noted that, as expected, the credible intervals increase as the forecast horizon increases, reflecting the uncertainties associated with the seasonal climate models used to generate the forecasts. For multiple provinces, the predicted mean number of dengue cases is above the 15*^th^* percentile outbreak threshold (i.e. yellow and orange shaded regions). This suggests that he April to September transmission season may be above normal conditions in multiple areas across Vietnam and particulary in Binh Doung, Dong Nai and Ho Chi Minh, all of which are located in the southern part of the country, characterised by endemic transmission.

## Discussion

We introduced a probabilistic dengue early warning system based on a model superensemble, formulated using Earth observations, and driven by seasonal climate forecasts. The system is able to generate accurate probabilistic forecasts of dengue metrics that could guide policy- and decision-making processes. We demonstrate that using a model superensemble results in better forecasts than using individual models.

Deciding which predictive model is the best from a suite of competing models is not a straightforward task. Each model carries somewhat different information of the modelled processes. Here, we present a method for reconciling between-model disagreements while improving forecast accuracy. The combination of models into a superensemble helps offset individual model biases across time and space **(*Krishnamurti et al., 2016*)**. Superensembles were initially developed for climate modelling **(*Krishnamurti et al., 1999*)**, and have gradually gained popularity in disease modelling (see for example ***Yamana et al., 2016,2017; Johansson et al., 2019*)**.

Our novel dengue early warning system relies on probabilistic models to properly reflect forecast uncertainty, and to explicitly assign probabilities to outcomes **(*Held et al., 2017*)**. The system has been developed to aid policy- and decision-making processes in Vietnam with the guidance of key stakeholders in the Ministry of Health of Vietnam, the World Health Organization regional office, the Pasteur Institutes in Nha Trang and Ho Chi Minh, Province-level Ministries of Health, the Vietnamese National Institute of Hygiene and Epidemiology, and the Tay Nguyen Institute of Hygiene and Epidemiology. A range of stakeholder engagement workshops, face-to-face meetings with users, and surveys were conducted to tailor the system to the users’ needs. Our results demonstrate that our spatio-temporal superensemble could guide changes to the current practice in dengue control towards a more preventative approach allowing bespoke and targeted public health interventions, and a more efficient allocation of scarce resources.

We demonstrate that the superensemble outperforms the predictive ability of any individual probabilistic model from a suite of top performing models in line with previous research **(*Krishnamurti et al., 1999; Yamana et al., 2016,2017; Johansson et al., 2019*)**. Model performance was assessed using a range of proper verification metrics across time and space to ensure the quality of our probabilistic predictions. To our knowledge, this is one of the first early warning systems informed by Earth observations to demonstrate skill for prospective year-round dengue prediction in a robust out-of-sample framework. It is also one of the first prototypes for routine dengue early warning at multiple time leads.

We found that the performance of the superensemble varied with geographic location, forecast horizon, and time of the year. The system showed skill in predicting spatiotemporal variations in dengue cases and outbreak occurrence at forecast horizons of up to six months ahead. Predictions deteriorated, and uncertainty increased as the forecast horizon expanded, in line with previous research **(*Reich et al., 2016; Chen et al., 2018; Funk et al., 2019; Tompkins et al., 2019*)**. Forecasts improved, and credible intervals decreased as time progressed and dengue data increased likely due to an improvement in the associations learned by the superensemble. Forecast skill is lower for the onset and peak of the transmission season due to substantial interannual variation (Supplementary Figure 7).

Relative to a baseline seasonal predictive model (see Methods and Materials), the superensemble made, on average, more accurate predictions across most provinces, and for most of the year in agreement with previous studies **(*Lowe et al., 2016; Chen et al., 2018*)**. This observation, however, conflicts with the results obtained by ***Lauer et al*. (*2018*)** and ***Johansson et al*. (*2019*)** for whom climate-naïve models had better skill than seasonal-climate-informed ones. ***Johansson et al*. (*2019*)** noted that incorporating seasonal climate data into predictive models may increase model complexity at the expense of lower out-of-sample forecast skill.

**Figure 7.**
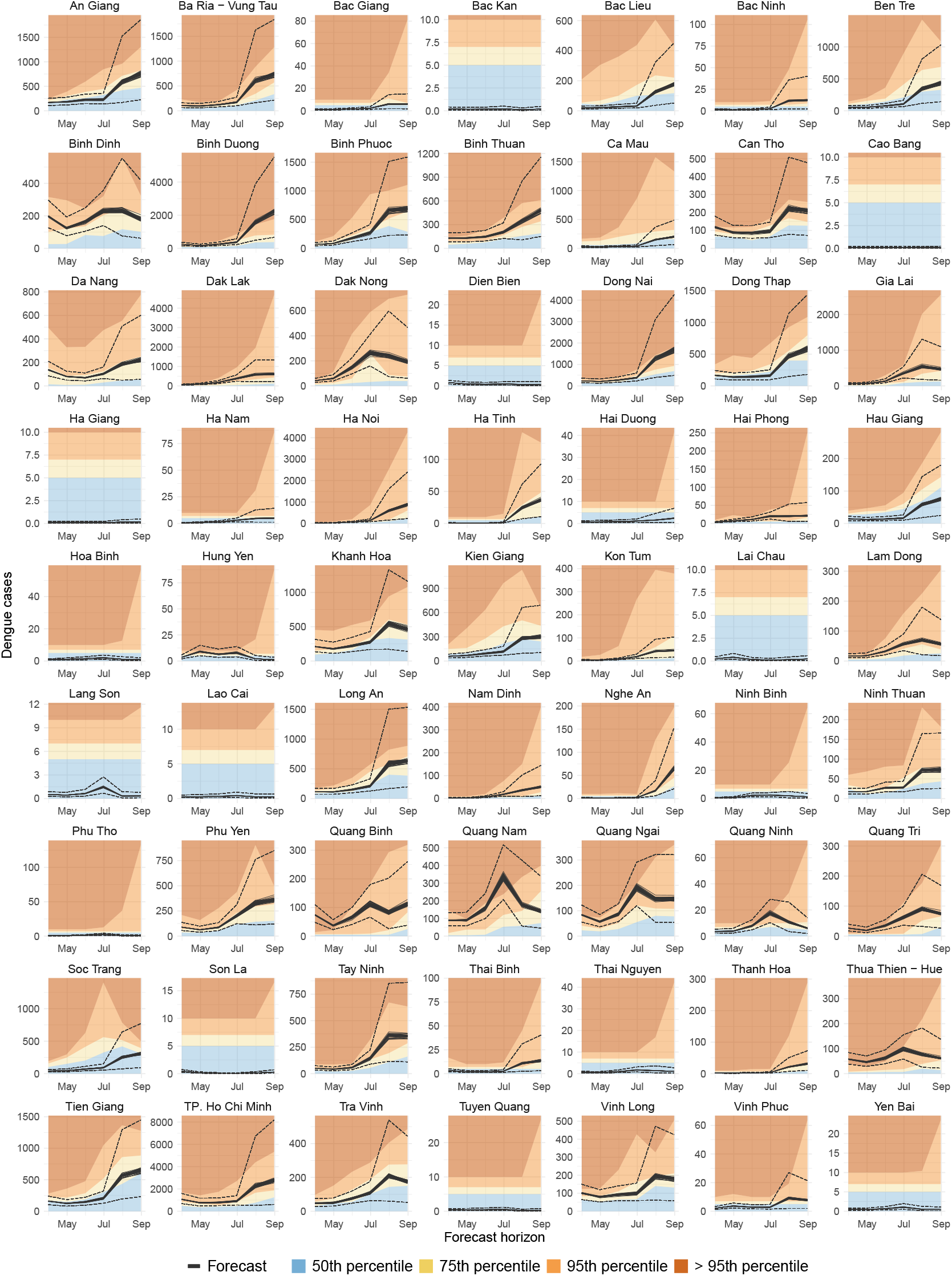
Predicted dengue cases for the period April to September 2020 for all Vietnamese provinces using a model superensemble. The forecast was issued on 5 April 2020. The *X* axis indicates the forecast horizon. The *Y* axis indicates the predicted number of dengue cases. Solid lines indicate the mean estimate for each of 42 forecast ensemble members. The dashed lines indicate the upper and lower bounds of the 95% credible intervals. The upper bound of the shaded areas indicates the month- and province-specific percentiles based on dengue data for the period August 2002 to March 2020.

Outbreaks are difficult to predict, even more so at forecast horizons of several months ahead. Nevertheless, our superensemble demonstrated skill for outbreak detection up to a lead time of six months, evaluated using proper scores over a suite of moving outbreak thresholds. One of the best performing outbreak thresholds was the endemic channel plus two standard deviations, computed using data from the previous five years. This method is currently in use across Vietnam (***Badurdeen et al., 2013; Runge Ranzinger et al., 2014*)** although with the difference of excluding outbreak years from the computation of the threshold. Following our results, however, we recommend the use of the 95% percentile as a threshold for outbreak detection as it gave better and more consistent results, likely due to its ability for detecting very large outbreaks. Recognizing the limitations of the province-level data, it is encouraging that our predictions are skillful in most provinces up to six months in advance.

In disease forecasting, each decision (e.g. to prevent or not to prevent an outbreak) has an associated cost that will lead to a benefit or a loss depending on the outcome. Decision-makers have the task of selecting the action that minimises potential losses. We used a simple cost-loss analysis **(*Thornes and Stephenson, 2001*)** to assess the relative economic value of the superensemble. Our figures are only illustrative. Still, they highlight that using a dengue early warning system has relative economic value compared to not using a forecast. The superensemble had considerable value across most provinces. However, in northern provinces, where dengue is essentially absent, the forecast is predicted to have limited relative economic value compared to always preparing for an outbreak or never preparing for it. The assessment of the relative economic value of the dengue early warning system may help stakeholders justify public investment in the development and generation of seasonal forecasts, or to help raise awareness of their potential value.

Although our proposed early warning system provides useful information for public health preparedness and response, it has some limitations worth mentioning. First, whilst our modelling framework incorporates important determinants of dengue occurrence such as climate and urbanisation, it does not explicitly incorporates, at this stage, the potential effects of other important determinants of disease such as the deployment of mosquito control interventions, vector indices, serotype-specific circulation, herd-immunity, and the mobility of people and goods all of which may lead to significant changes in the level of risk experienced locally **(*Reiter, 2001*)**. Stakeholders in Vietnam have recently started collecting supporting data on vector indices and dengue virus serotypes. However, it is noted that there is a lack of publicly available, continuous, and long-term data sets that could be used to inform modelling efforts. In this study, we account for some of the variation that might be attributed to these factors by using spatio-temporal random effects in each of the models included in the superensemble. Future developments of the system may incorporate some of these factors if data are made available. Second, the quality and consistency of the dengue data are affected by the limited confirmation of suspected cases through laboratory diagnostic test, leading to large uncertainties that are difficult to quantify. Third, our computations of dengue risk do not take into consideration uncertainties due to the potential under- or misreporting of dengue cases. Consequently, our model superensemble forecasts may underestimate the real number of cases occurring at any given time.

### Conclusion

We have demonstrated that a probabilistic dengue early warning model, formulated using Earth observations and driven by seasonal climate forecasts, to produce probabilistic predictions of exceeding pre-defined outbreak-detection thresholds. A theoretical cost-loss analysis showed that the system has relative economic value for a range of cost-loss ratios across most of the country, indicating that such a system may be useful for decision-making processes. The dengue forecasting system presented here has been rolled out across Vietnam and could be tailored for other dengue-endemic countries. Further work may include investigating the feasibility of producing probabilistic forecasts with sufficient skill at the district or commune levels.

## Methods and Materials

### Dengue surveillanc0e data

Monthly dengue cases were obtained from the Vietnamese Ministry of Health. Data were retrieved for the period August 2002 to December 2019 at the province level (*n* = 63). Data comprised suspected and confirmed dengue cases although there was no indication as to how many cases fell within each category. The data set did not contain serotype-specific information.

### Historical Earth observation data

Minimum, maximum and mean air temperature at 2 metres above ground (°C) were derived from MODIS daily L3 global land surface temperature products (***Wan et al., 2015*)** with a spatial resolution of 1 km^2^. Precipitation amount per day (mm day^−1^), was initially retrieved from from the Tropical Rainfall Measurement Mission (***Hijmans, 2011*)** at a spatial resolution of 25 km^2^ up to April 2014. After April 2014, precipitation data were obtained from the Global Precipitation Mission **(*Huffman et al., 2019*)** at a spatial resolution of 10 km^2^. Daytime specific surface humidity (kg kg^−1^) was calculated using the daytime MODIS L2 water vapour near infrared MOD 5 products **(*Gao, 2015*)** with a spatial resolution of 1 km^2^. Average daily wind speed at 10 metres above ground (m s^−1^) was retrieved from the European Centre for Medium-range Weather Forecasts (ECMWF) ERA-5 reanalysis **(*Copernicus, 2019*)** for the period 2002-2011, at a spatial resolution of 31 km^2^. After 2011, wind speed data were obtained from the NOAA Climate Forecast System **(*Saha et al., 2014*)** at a 20 km^2^ resolution. Monthly sea surface temperature anomalies for the Niño region 3.4 (5°S-5°N, and 170°–120°W) were obtained from the NOAA Center for Weather and Climate Prediction, Climate Prediction Center **(*NOAA, 2020*)** for the period 2002 to 2020. Population weighted monthly averages for each climate variable were calculated for each province.

Population data from the Worldpop project **(*WorldPop, 2018*)** at a 100 m^2^ spatial resolution was used to calculate annual gridded weights for each province. At the time of processing, Worldpop data were only available for the years, 2009, 2010, 2015, 2020. Intervening years were generated using linear interpolation.

### Historical demographic and land-cover data

Total population per province were retrieved from the Socioeconomic Data and Applications Center (SEDAC) Gridded Population of the World project version 4.11 **(*CIESIN, 2019*)** at a 1 km^2^ resolution, 5-yearly for the period 2000 to 2020. Intervening years were generated using linear interpolation. The percentage of urban, peri-urban and rural land-cover per province for the period 2002-2019 was derived from the ESA CCI Land-cover project **(*ESA, 2017; Copernicus, 2019*)** which describes the land surface into 22 classes at a spatial resolution of 0.00277 degrees.

### Seasonal climate forecasts

Seasonal climate forecasts of minimum temperature, maximum temperature, daily precipitation, specific humidity, wind speed and sea surface temperature anomalies for the Nino region 3.4 were obtained from the UK Met Office Global Seasonal Forecasting System version 5 (GloSea5) **(*MacLachlan et al., 2015; Scaife et al., 2014*)**. GloSea5 comprises 42 ensemble members built around a high resolution climate prediction model (HadGEM3). Ensemble members differ due to small stochastic physics perturbations provided by the Stochastic Kinetic Energy Backscatter v2 **(*Bowler et al., 2009*)**. GloSea5 has a resolution of 0.83 degrees in latitude and 0.56 degrees in longitude for the atmosphere, and 0.25 x 0.25 degrees for the ocean. GloSea5 has two major components, the forecast itself and the associated hindcasts or *historical re-forecasts*, which are used for calibration and skill assessment.

### Seasonal climate hindcasts

Hindcast data (i.e. historical forecasts) were retrieved from the Copernicus Climate Data Store **(*Copernicus, 2019*)** at monthly time steps for each of the 28 ensemble members. Data were obtained using the GloSea5 system (***MacLachlan et al., 2015; Scaife et al., 2014*)** across a forecast horizon of one to six months ahead for the period January 2012 to December 2016. At the time of the computations, data for the period May to October 2016 were unavailable.

### Modelling approach

Let *Y_i,t_* be the number of dengue cases for province *i* = 1,…, *I* at time *t* =1, ⋯, *T* be modelled using Bayesian generalised linear mixed models (GLMM). Models were fitted using a negative binomial specification to account for potential over-dispersion in the data. The general algebraic definition of the models is given by:

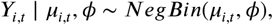

where *μ_i,t_* is the expected number of dengue cases for province *i* and time *t*, and *ϕ* > 0 is the negative binomial dispersion parameter. A logarithmic link function of the expected number of cases is modelled as:

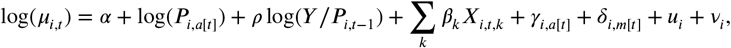

where *α* corresponds to the intercept; log(*P_i_*,*_a_*_[_*_t_*_]_) denotes the logarithm of the population at risk for province *i* and year *a*[*t*], included as an offset to adjust case counts by population; *ρ* is the auto-regressive coefficient; log(*Y/P_i,t−_*_1_) is the logarithm of the observed dengue incidence rate in the previous month with regression coefficient *ρ*; *X* is a matrix of *k* seasonal Earth observation (meteorological and land-cover type) explanatory variables with regression coefficients *β*. Longterm trends are modelled using province-specific unstructured random effects for each year (*γ_i,a_*_[_*_t_*_]_). Seasonality is accounted for by using province-specific structured random effects for each calendar month (*δ_i,m_*_[_*_t_*_]_) with first order auto-regressive prior to allow each month to depend on the previous one. Unknown confounding factors, such as interventions and spatial dependency structures representing, for example, human mobility, were incorporated using structured (*v_i_*) and unstructured (*u_i_*) random effects for each province *i*. Spatial random effects were specified using a Besag-York-Mollie model **(*Besag et al., 1991*)** which incorporates a spatial effect with a Gaussian exchangeable prior to account for unstructured variation, and a spatial effect with an intrinsic conditional auto-regressive prior to account for spatially-structured variability. Delayed effects of meteorological factors on dengue were accounted for by incorporating the moving average of temperature, precipitation, specific humidity, and diurnal temperature range (DTR) all of them lagged zero to two months, and sea surface temperature anomalies in the Niño region 3.4 lagged three months based on previous research (e.g. **(*Petrova et al., 2019; Colón-González et al., 2018a; Lowe et al., 2018*)** and exploratory analyses. No delayed effects were considered for wind speed. Flat priors were set to regression coefficients (*α,ρ,β*) and penalising complexity priors were assumed for both the dispersion parameters and the precision for all random effects **(*Simpson et al., 2017*)**. Models were fitted in R version 3.6.1 using the INLA package **(*Rue et al., 2009*)**. The relevant R code is available at https://github.com/FelipeJColon/paper_dengue_superensemble.

### Model selection

The best subset of seasonal climate predictors leading to the lowest observed-prediction discrepancies for a given model was obtained using an expanding window time series cross-validation (TSCV) algorithm **(*Hyndman and Athanasopoulos, 2014*)**. Land-cover variables were included in all models as they varied annually. We iteratively fitted all possible models containing one seasonal climate predictor at the time, then two seasonal climate predictors, and so on, until all seasonal climate variables were included in a full model (***Colón-González et al., 2018a*)**. The predictive ability of each model was evaluated using the continuous rank probability score (CRPS), root mean squared error (RMSE), mean absolute error (MAE) deviance information criterion (DIC) and the Watanabe-Akaike information criterion (WAIC). Verification metrics were computed in R using the SpecsVerification and ModelMetrics packages **(*Siegert, 2017; Hunt, 2018*)**.

TSCV was implemented using an expanding window approach dividing the data set into multiple training and testing sets. The initial training set comprised data from August 2002 to December 2006. Each time step (*k*), a further month of data was added to the training set until the training set contained *n−*6 observations. The testing set comprised the climate hindcast data for the six months immediately after the last observation in the training set for each geographical area. Seasonal climate hindcast data comprised 28 ensemble members.

We calculated the mean CRPS, RMSE, MAE, DIC and WAIC for each model specification across all time steps and ensemble members. The best two performing models for each verification metric were selected to create a superensemble.

### Accounting for autocorrelation in disease transmission

The number of dengue cases occurring at time *t* is directly dependent on the number of cases that occurred in the recent past. Previous research suggests that including the logarithm of the number of cases in the previous month log(*Y_t−_*_1_) helps accounting for such temporal correlation in disease transmission **(*Imai et al., 2015*)**. Additionally, incorporating log(*Y_t−_*_1_) as a covariate improves the predictive ability of seasonal-climate-informed disease models by reducing residual dispersion **(*Imai et al., 2015*)**. One complication of accounting for the number of cases in the previous month in an operational system is that log(*Y_t−_*_1_) in the temporal window of the forecast can only be known up to time *t* + 1. More specifically, to generate dengue forecasts one month ahead, log(*Y_i_*,*_t−_*_1_) for time *t* + 1 corresponds to the logarithm of the number of dengue cases at time t. To generate the forecasts two months ahead we first fitted a climate naïve model with the following specification:

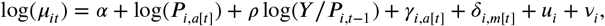

with *α* as the intercept; log(*P_i_*,*_a_*_[_*_t_*_]_) as the population at risk in province *i* at time *a*[*t*], included as an offset; log(*Y/P_i,t_*_−1_) is the logarithm of the observed dengue incidence rate in the previous month with regression coefficient *ρ*; *γ_i_,_a[t]_* as province-specific unstructured random effects; *δ_i,m_*_[_*_t_*_]_ as province-specific structured random effects with an AR1 auto-correlation term; and *u_i_* and *ν_i_* as province-specific structured and unstructured random effects. We then predicted the number of dengue cases for time *t* + 1. The logarithm of the predicted number of cases at time *t* + 1 was then used as log(*Y_i_,_t−_*_1_) for time *t* + 2. We repeated these steps for each lead time in the forecast. Given the high number of zero counts in the set, we used the logarithm of the number of dengue cases lagged 1 month plus one.

### Baseline model

A baseline model was developed for comparison purposes. The algebraic definition of the model is given by:

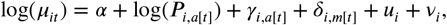

Where *α* is the intercept; log(*P_i,a_*_[_*<u>_t_</u>*_]_) is the population at risk in province *i* at time *a*[*t*], included as an offset; *γ_i,a_*_[_*_t_*_]_ are province-specific unstructured random effects; *δ_i_*,*_m_*_[_*_t_*_]_ are province-specific structured random effects with an AR1 auto-correlation term; and *u_i_* and *ν_i_* as province-specific structured and unstructured random effects.

### Model superensemble

Given a number of competing models, we generated a model superensemble **(*Yamana et al., 2016*)** using Bayesian model averaging (BMA) implemented in the INLABMA package **(*Bivand et al., 2015*)**. BMA relies on the weighted sum of the conditional marginals obtained from a group of competing models **(*Gómez-Rubio et al., 2019*)**. The posterior distribution given data *D* is estimated as follows:

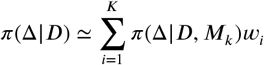

with Δ as the quantity of interest; *M_k_* indicates the *M*_1_…*M_K_* competing models that will form the superensemble; and the weights *w_i_* computed based on the maximum likelihood (ML) and deviance information criterion (DIC) of the competing models. The weights *w_i_* were calculated as follows:

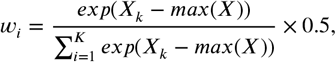

where *X* is a vector of ML or DIC values for each of the competing models *k*. The vector of ML and DIC weights were multiplied by 0.5 so that the resulting superensemble weights would sum to one, and then added together into a single vector *w_i_*. A new set of superensemble weights were computed each time a forecast was issued using training data from all previous years.

The predictive ability of the superensemble was evaluated using the CRPS, RMSE and MAE. In addition, we evaluated the skill of the obtained forecasts using the continuous rank probability skill score (CRPSS). CRPSS is defined as:

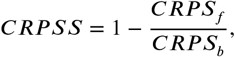

where *CRPS_f_* is the continuous rank probability score (CRPS) of the forecast and *CRPS_b_* is the CRPS of a baseline model used as a reference **(*Bradley and Schwartz, 2011*)**.

### Outbreak detection evaluation

The skill of the model for detecting dengue outbreaks was evaluated using the Brier **(*Brier, 1950*)** and logarithmic scores **(*Good, 1952*)** both of which are strictly proper scoring rules based on probability densities. The Brier score was calculated as follows:

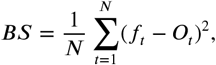

where *N* is the number of predictions; *f_t_* is the forecast probability that an outbreak may happen; and *O_t_* takes the value of one if there was an outbreak, or zero if there was no outbreak. The logarithmic score was calculated as follows:

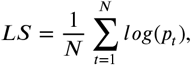

where *p_t_* denotes the probability assigned to the observed outcome over *N* predictions. Scores were computed in R using the scoring package **(*Merkle and Steyvers, 2013*)**.

### Relative economic value

Unlike skill, the relative economic value (*V*) of the model superensemble depends on requirements set by the user **(*Thornes and Stephenson, 2001*)**. Typically, *V* is evaluated in monetary terms and is particularly useful when the probability of occurrence of an adverse event (e.g. a major outbreak) is known. If the probability of occurrence of an outbreak is greater than the ratio of the cost of taking preventive action divided by the loss incurred by not taking action (C/L ratio) and an outbreak occurs, then it will pay off to take action. If the probability of occurrence of an outbreak is lower than the C/L ratio, then it does not pay off to take preventive action. If the probability is equal to the C/L ratio, it does not matter if action is taken or not.

*V* was estimated for a range of theoretical C/L ratios following **(*Thornes and Stephenson, 2001*** by comparing the mean cost of using the forecasting system for outbreak detection compared to the mean expense incurred by either never preventing outbreaks or, on the contrary, taking preventive action every month of the year. *V* takes a value of one if the forecast is perfect, and a value of zero if it is no better than the default action plan. If *V* is negative, it indicates that the forecast is so poor that it would be more cost effective not to use it. *V* is represented as follows:

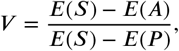

where *E*(*S*) is the expense incurred by taking preventive action each month of the year or the losses incurred by no taking action at all even when an outbreak occurred, whichever is the cheapest method when not using the forecast; *E*(*A*) represents the total cost of the forecast calculated as the cost of type 1 (false positive) and type 2 (false negative) errors plus the cost of acting when the forecast was correct; and *E*(*P*) indicates the cost incurred with a perfect forecast (i.e action is taken only when there is an outbreak).

## Data Availability

At the time of the submission, data cannot be released to third parties due to confidentiality agreements between the members of the consortium and theGeneral Department of Preventive Medicine, Vietnam. All relevant code is available in a GitHub repository.

https://github.com/FelipeJColon/paper_dengue_superensemble

## Funding

UK Space Agency Dengue forecasting MOdel Satellite-based System (D-MOSS)

- Felipe J Colón-González
- Leonardo Soares Bastos
- Barbara Hofmann
- Alison Hopkin
- Quillon Harpham
- Tom Crocker
- Rosanna Amato
- Iacopo Ferrario
- Francesca Moschini
- Samuel James
- Sajni Malde
- Eleanor Ainscoe
- Mark Harrison
- Gina Tsarouchi
- Darren Lumbroso
- Oliver Brady
- Rachel Lowe

The funders had no role in study design, data collection and interpretation, or the decision to submit the work for publication.

## Acknowledgments

We would like to thank Dr Satoko Otsu, and Dr Trang Cong Dai Dai from the World Health Organization Office in Vietnam, and Mr Dao Khanh Tung from the United Nations Development Programme Office in Vietnam for the helpful discussions and support for the development of this project. We would also like to acknowledge the support and insights of the Pasteur Institute Ho Chi Minh City, the Pasteur Institute Nha Trang, the Institute of Hygiene and Epidemiology Tay Nguyen (TIHE), and the National Institute of Hygiene and Epidemiology (NIHE).

## Appendix

Supplementary Figures

**Appendix 1 Figure 1.**
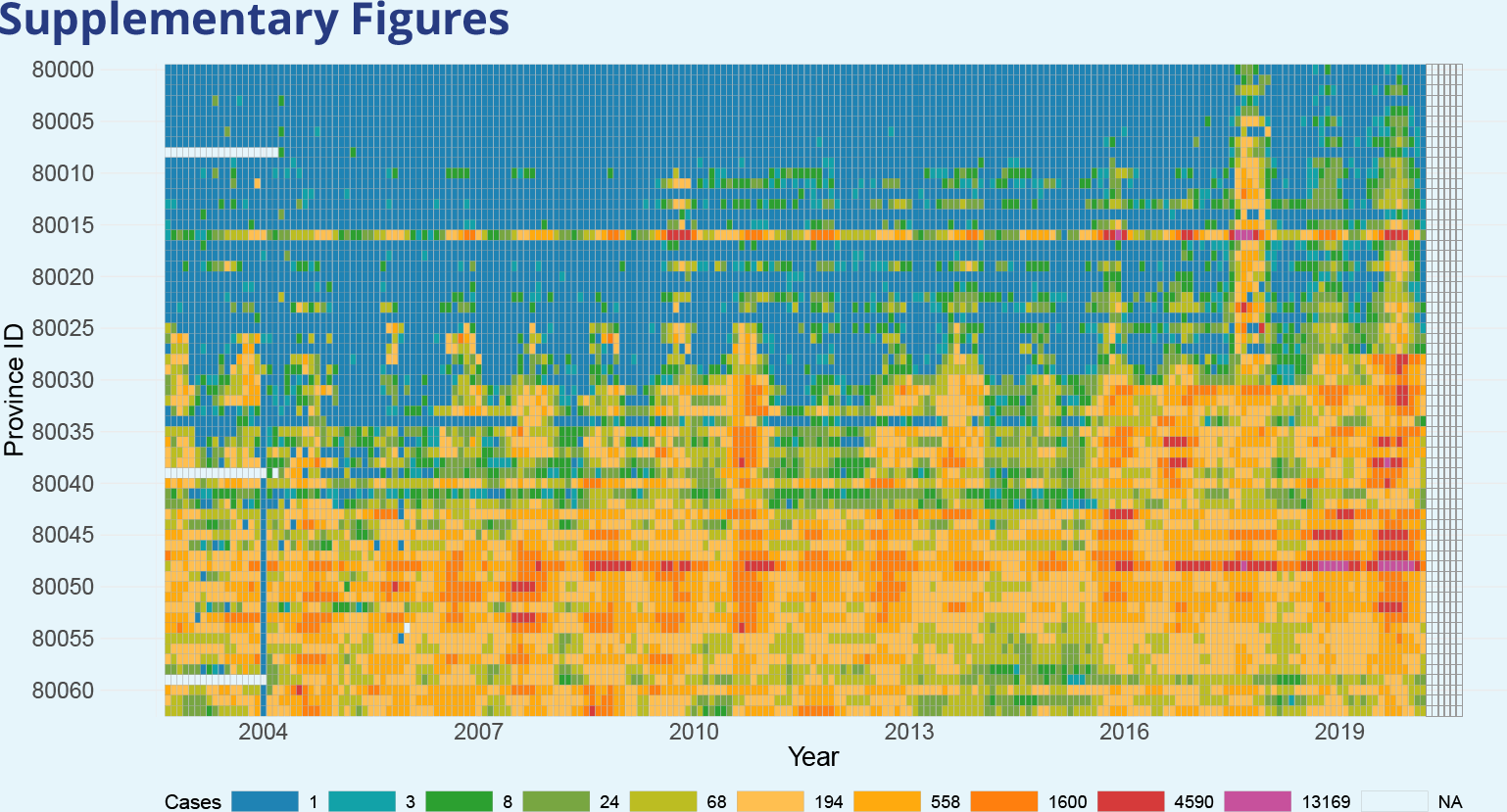
Time series of monthly dengue cases from the 63 provinces in Vietnam (Aug 2002 to March 2020). Provinces are ordered from north (top) to south (bottom) according to the latitude coordinates of their centroid. White boxes indicate missing data.

**Appendix 1 Figure 2.**
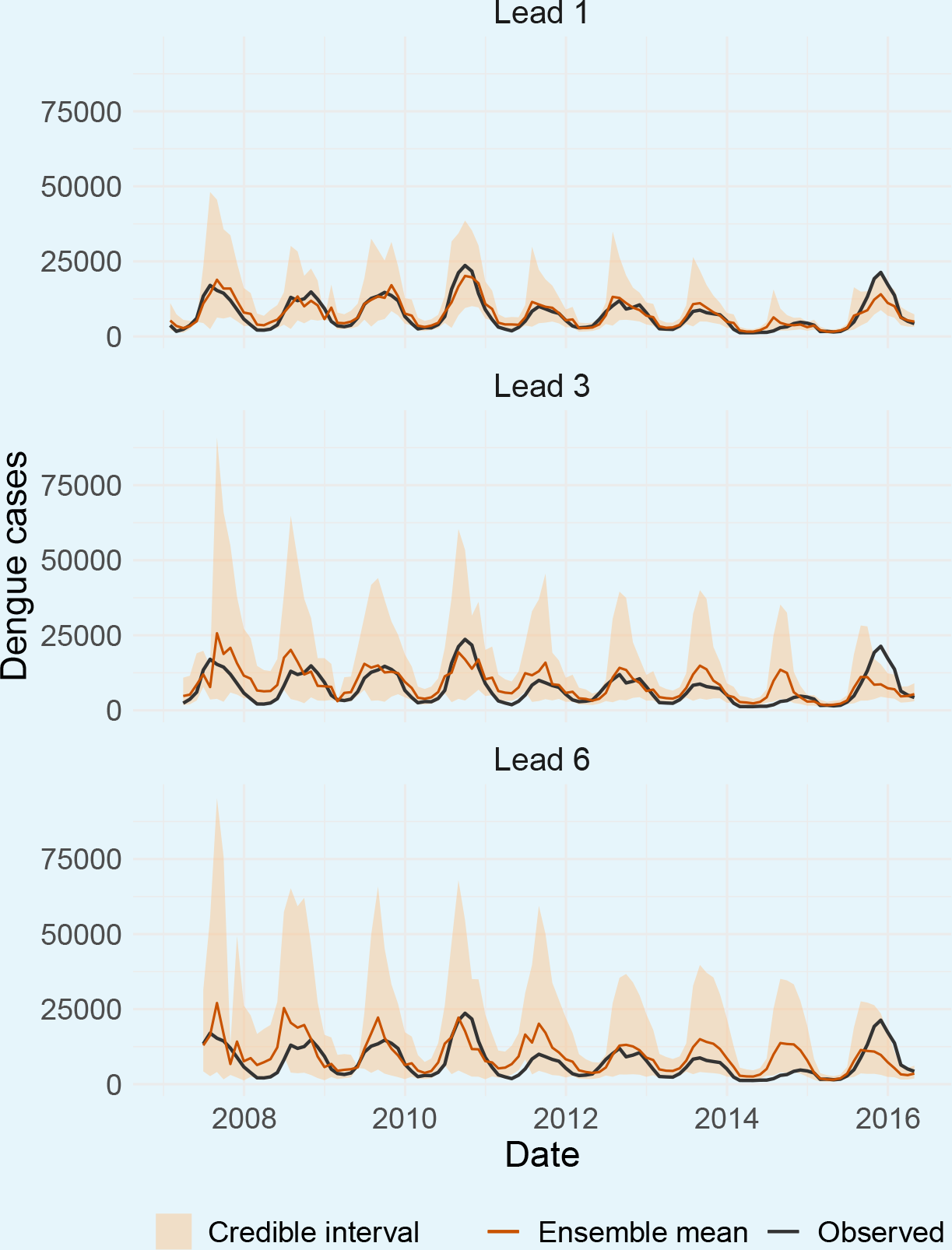
Observed (solid black lines) and predicted (solid red lines) dengue cases across Vietnam aggregated at the national level. Shaded areas represent the 95% credible interval. Predictions are shown for the forecast horizons of one (top), three (middle), and six (bottom) months ahead. Data corresponds to the period January 2007 to December 2016.

**Appendix 1 Figure 3.**
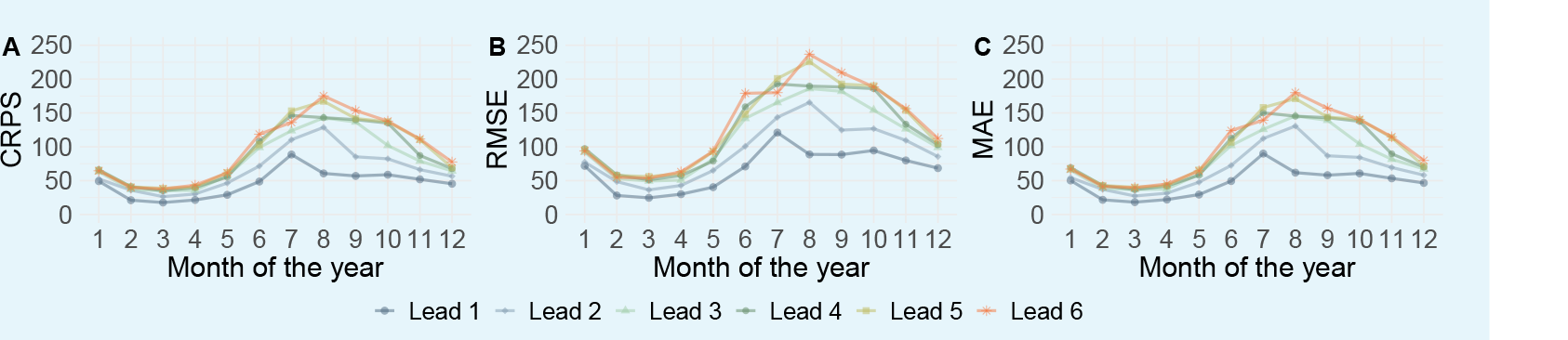
Continuous rank probability score (A), root mean squared error (B), and mean absolute error (C) of the model superensemble by month and forecast horizon.

**Appendix 1 Figure 4.**
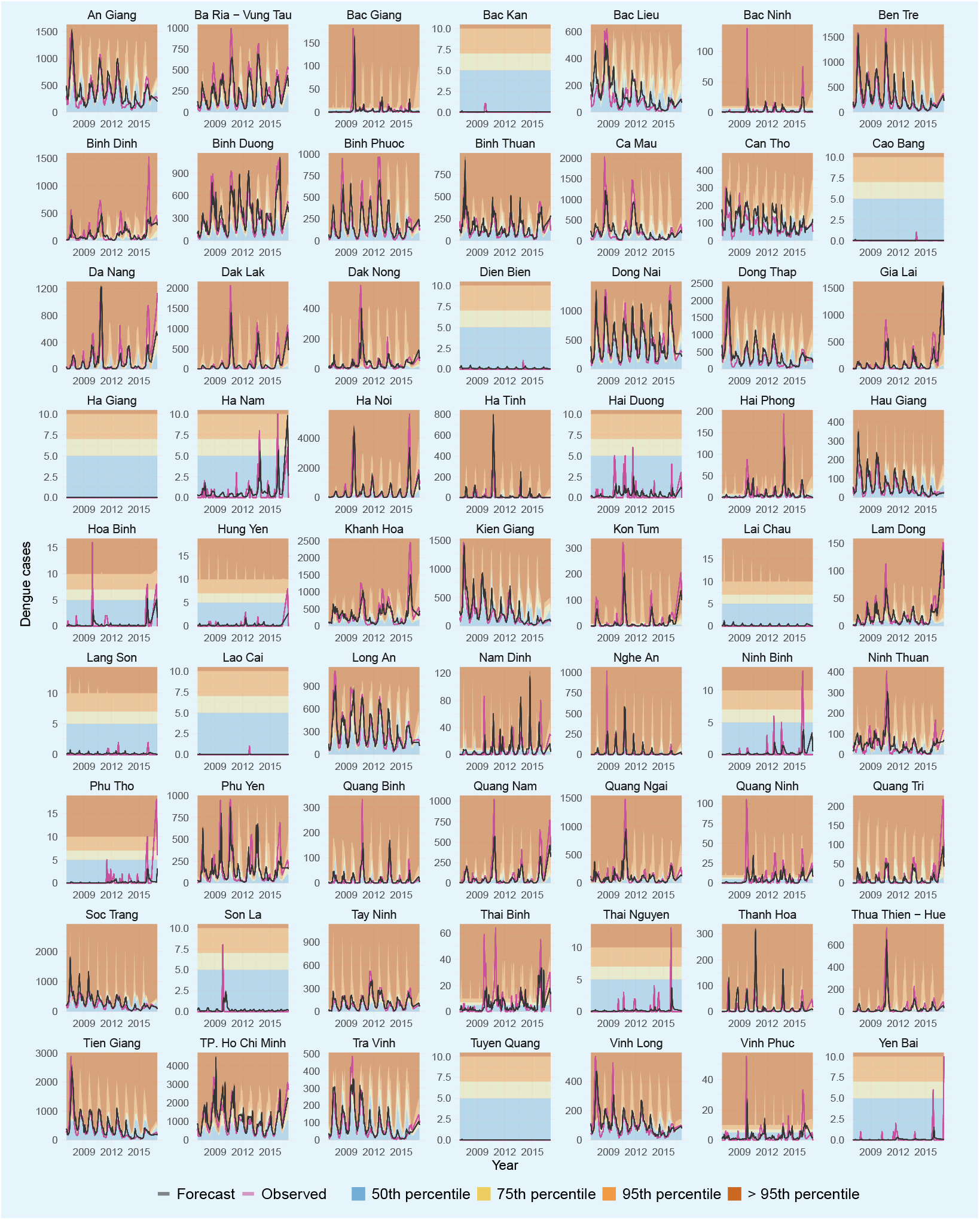
Observed (solid black lines) and predicted (solid blue lines) dengue cases across 63 Vietnamese provinces computed one month ahead. Data corresponds to the period January 2007 to December 2016. Shaded areas represent a range of month- and province-specific percentiles to guide public health decision-making.

**Appendix 1 Figure 5.**
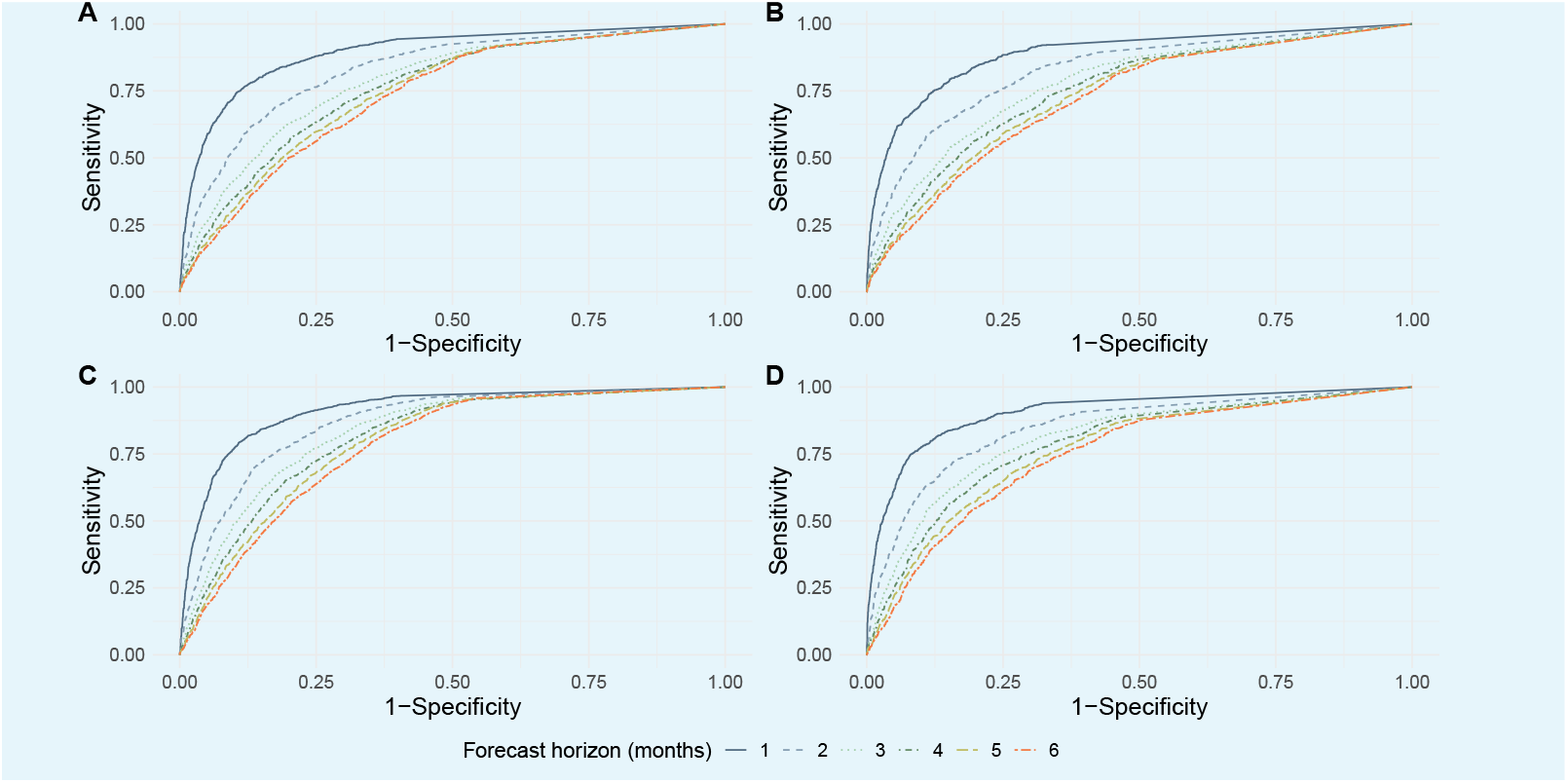
Area under the curve (AUC) for different probabilities of exceeding four different moving outbreak thresholds: one (A) and two (B) standard deviations above the mean, 75*^th^* (C) and 95*^th^* (D) percentiles.

**Appendix 1 Figure 6.**
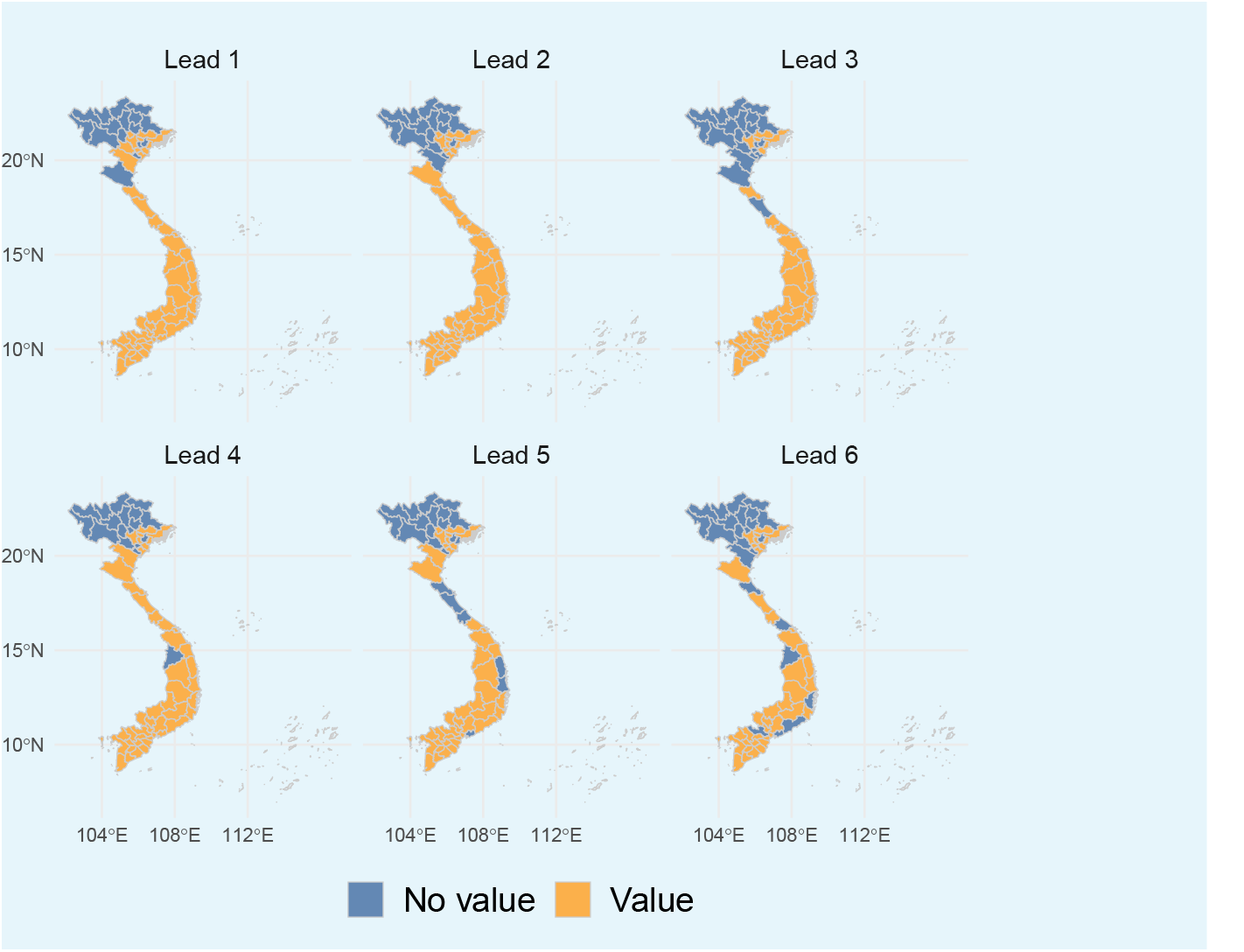
Spatial variation of the relative economic value of the model superensemble over the forecast horizon of one to six months. Orange shaded areas indicate provinces where there is relative economic value (based on a range of theoretical cost-loss ratios and outbreak thresholds). Blue shaded areas indicate provinces where the superensemble had no relative economic value.

**Appendix 1 Figure 7.**
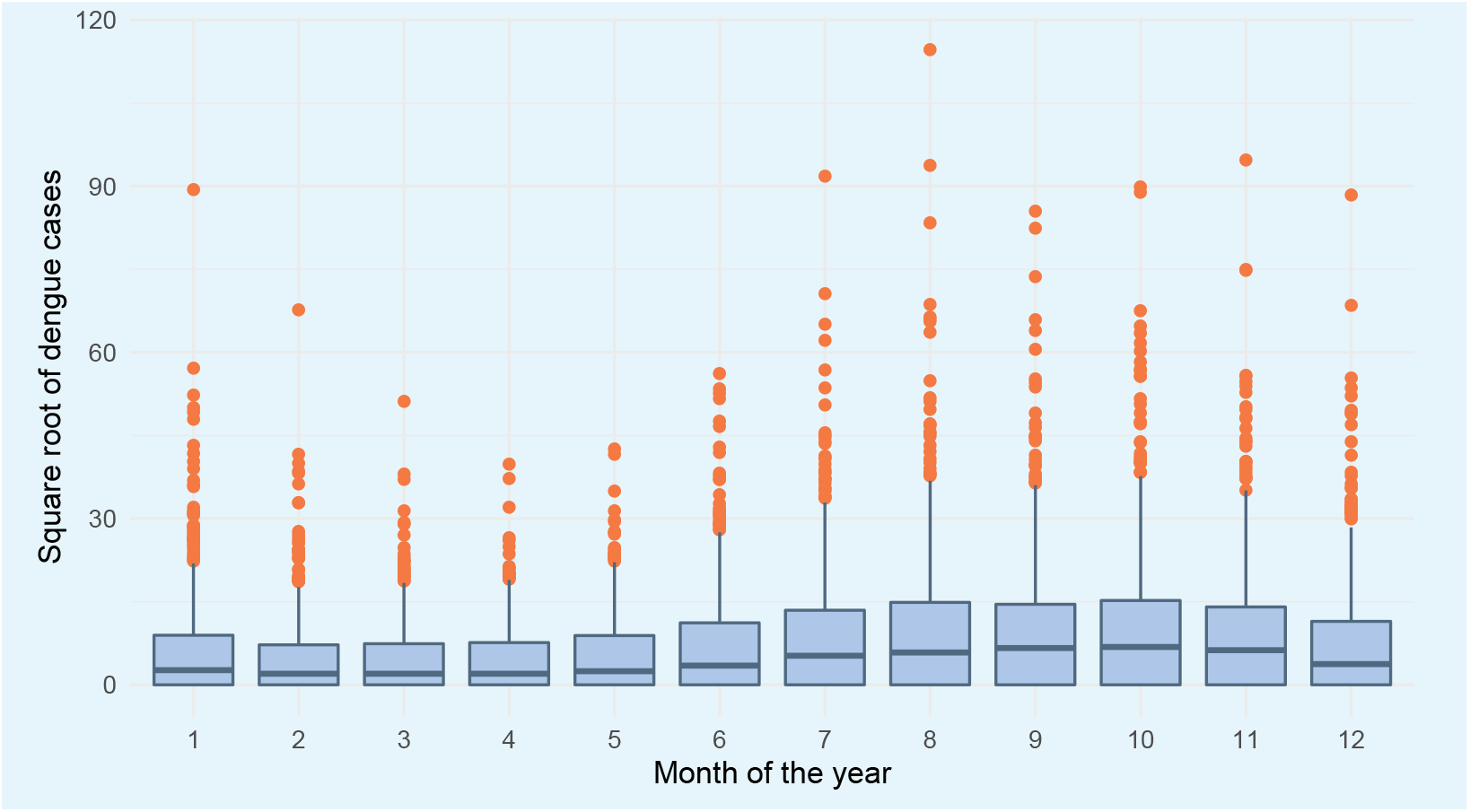
Month-specific variability in dengue cases across Vietnam. The *X* axis indicates the month of the year. The *Y* axis indicates increases in the number of dengue cases (square root transformed). The upper and lower limits of each box represent the inter-quartile range of the distribution of dengue cases for each month. The middle solid line indicates the median value. The upper and lower whiskers indicate the maximum and minimum values of the dengue case distribution (excluding outliers which are indicated in orange circles). Outliers are values beyond ±1.5 times the inter-quartile range.

